# Five-year Efficacy and Safety of TiNO-Coated Stents Versus Drug-Eluting Stents in Acute Coronary Syndrome. A meta-analysis

**DOI:** 10.1101/2023.08.31.23294251

**Authors:** Frederic C. Daoud, Bogdan Catargi, Pasi P. Karjalainen, Edouard Gerbaud

**Affiliations:** Endocrinology-Metabolic Diseases, Hôpital Saint-André, Bordeaux University, 33000 Bordeaux, France; (F.D.); (B.C.); Cardiac Unit, Heart and Lung Center, Helsinki University Hospital, Helsinki University, 00280 Helsinki, Finland; (P.K.); Cardiology Intensive Care Unit and Interventional Cardiology, Hôpital Cardiologique du Haut-Lévêque, 33604 Pessac, France; (E.G.); Bordeaux Cardio-Thoracic Research Centre, U1045, Bordeaux University, 33076 Bordeaux, France

**Keywords:** acute coronary syndrome, non-drug-eluting titanium-nitride-oxide coated stents (TiNOS), drug-eluting stents, 5-year follow-up, safety, efficacy, systematic literature review, meta-analysis

## Abstract

(1) Background: Percutaneous Coronary Interventions (PCI) in patients with acute coronary syndrome (ACS) are performed with titanium-nitride-oxide-coated stents (TiNOS) or drug-eluting stents (DES). This prospective systematic literature review (SLR) of prospective, randomized, controlled trials (RCTs) showed TiNOS is non-inferior to DES in major adverse cardiac events (MACE) rates and presents a lower risk of recurrent myocardial infarction (MI) at 1-year follow-up. (2) Methods: The previously described SLR and meta-analysis protocol, per PRISMA, Cochrane methods, and GRADE, applied here to 5-year follow-up outcomes. (3) Results: Three RCTs were eligible, comprising 1,620 patients with TiNOS vs. 1,123 with DES. The pooled risk ratios (RRs) and 95% confidence intervals were: MACE 0.82 [0.68, 0.99], MI 0.58 [0.44, 0.78], cardiac death (CD) 0.46 [0.28, 0.76], ischemia-driven target lesion re-vascularization (TLR) 1.03 [0.79, 1.33], probable or definite stent thrombosis (ST) 0.32 [0.21, 0.59], and all-cause mortality (TD) 0.84 [0.63, 1.12]. Evidence certainty was high in MACE, CD, MI, and ST. Certainty was moderate in TLR and TD. (4) Conclusions: TiNOS in ACS at 5-year follow-up appears safer than DES and equally efficacious. Stratified pooled outcomes by clinical presentation would facilitate clinical validity benchmarking and generalization to target populations.

## 1. Introduction

Acute Coronary Syndrome (ACS) designates sudden myocardial ischemia. It comprises three clinical presentations: ST-segment elevation myocardial infarction (STEMI), non-ST-segment elevated myocardial infarction (NSTEMI), and unstable angina pectoris (UAP) [1]. A previous prospective Systematic Literature Review (SLR) and meta-analysis of prospective Randomized Con-trolled Trials (RCT) compared the 1-year follow-up clinical outcomes after Percutaneous Coronary Interventions (PCI) using non-drug-eluting Titanium-Nitride-Oxide-coated Stents (TiNOS) to treat patients with ACS [2]. The pooled Risk Ratios (RRs) showed that TiNOS was non-inferior to DES in terms of device-oriented Major Adverse Cardiac Events (MACE) rates and presented a lower risk of recurrent myocardial infarction (MI) at 1-year follow-up.

## 2. Materials and Methods

This SLR was conducted according to the previous prospective protocol (PROSPERO CRD4201809062) that planned 5-year follow-up outcomes analysis upon patient follow-up completion [3].

The PICO framework was used to define the research question [4]. The SLR was conducted according to the principles described in the Cochrane Handbook and the “Preferred Reporting Items for Systematic Reviews and Meta-Analyses” (PRISMA), and certainty of evidence was assessed according to the “Grading of Recommendations Assessment, Development, and Evaluation” (GRADE) [5–12]. The primary endpoint was device-oriented Major Adverse Cardiac Events (MACE). The secondary endpoints were Cardiac Death (CD), recurrent non-fatal Myocardial Infarction (MI), ischemia-driven Target Lesion Revascularization (TLR), Probable or definite Stent Thrombosis (ST), and all-cause mortality (“Total Death” TD). The endpoint definitions were those of the Academic Research Consortium (ARC-2) [13]. The detailed SLR and meta-analytic methods applied with 5-year outcomes were similar to those used with 1-year outcomes and previously published, except for the updates below [2,3].

F.D. and B.C. were the primary reviewers of 5-year outcomes, E.G. adju-dicated disagreements, and P.K. obtained checked data in case of uncertainty.

The 5-year follow-up outcomes meta-analysis was computed in the 2023 versions of the same software packages as used with 1-year follow-up: RevMan, Harbord tests in STATA (version 17, StataCorp LP, College Station, TX, USA) using the metan and metabias packages, and GRADE analysis was performed in GRADEpro GDT software 2023 online (https://gradepro.org) [14,15]. Pubmed, Scopus, the Cochrane Library, and Web of Science (WoS) electronic databases were queried on July 5, 2023, with the same search string as previously [2]. The interpretation of the string by each database’s search engine is in Appendix A.

To avoid deriving conclusions about patients with chronic coronary syndrome (CCS) based on data from patients presenting with ACS, RCT eligibility was narrowed down to trials that either included patients with ACS only or included a mix of patients with ACS and CCS providing baseline and outcomes data in ACS and CCS were reported separately.

To improve the clinical intuitiveness of pooled RR imprecision rating, GRADE’s 2011 guideline 6 was replaced by the minimally contextualized approach (guidance 34, 2022) [11,12]. An outcome’s pooled RR was defined as imprecise if its 95% Confidence interval (CI) crossed threshold lines of appre-ciable harm, benefit, or futility, depending on the situation. The lower and upper RR thresholds were 0.9 and symmetrically 1.11 (i.e., 1/0.9). A relative risk reduction (RRR) of 10%, the “futility threshold” (lower threshold RR = 1-RRR) in the Cochrane review of DES vs. BMS in ACS defined the thresholds [16]. In MACE and TLR, where DES was assumed to be superior to TiNOS, the RR of events in TiNOS over DES was tested for non-inferiority, and the CI was rated seriously imprecise if it crossed the upper threshold. Safety outcomes RRs (CD, MI, TD, ST) underwent a two-sided superiority test with the usual null hypothesis of no difference between the event rates of the compared treatments. A significant RR’s CI was rated seriously imprecise if it crossed the nearest threshold. Non-significant RR CIs crossing one or both thresholds were rated seriously imprecise.

## 3. Results

### 3.1. Study Identification and Selection

Ninety-four records were identified, and nine publications with first-hand data about three RCTs were found eligible for pooling after selection (PRISMA flowchart in Figure 1) [5]. The 49 excluded records did not meet the PICO specifications or were review articles. One RCT comparing TiNOS to zotarolimus-eluting stents included 53% with CCS and 47% with ACS, but the study was excluded from the pooled analysis because it did not stratify the data [17,18].

**Figure 1.**
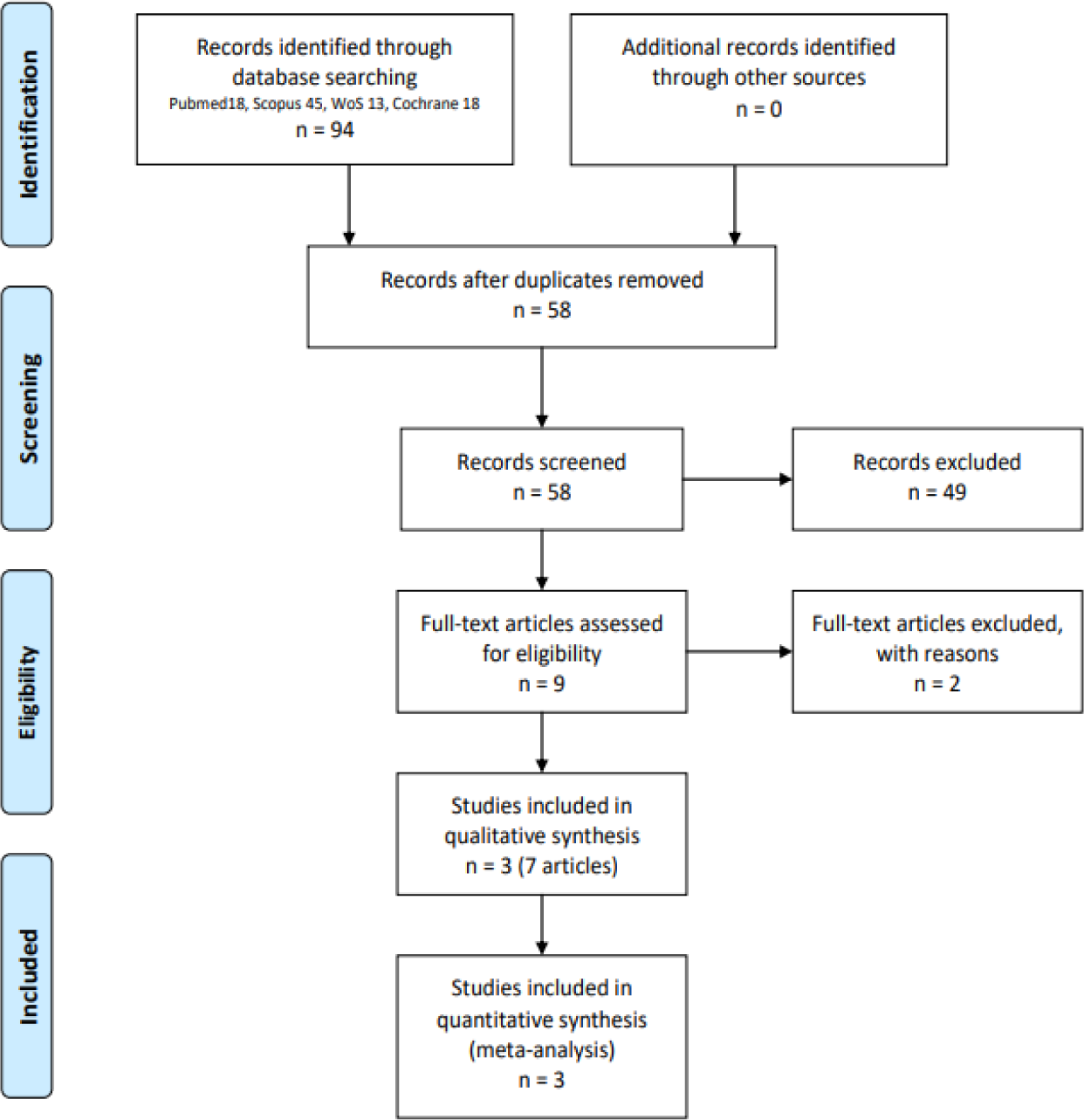
PRISMA flowchart.

### 3.2. Individual Study Characteristics

Table 1 summarizes the characteristics of the eligible RCTs and the re-ported raw data. The overall numbers and rates of patients lost to follow-up were 4.63% (67/1620) in the TiNOS arm and 4.36% (49/1123) in the DES arm. The pooled distribution of clinical presentation

ions was NSTEMI 48.2%, STEMI 43.2%, and UAP 8.6%.

**Table 1.**
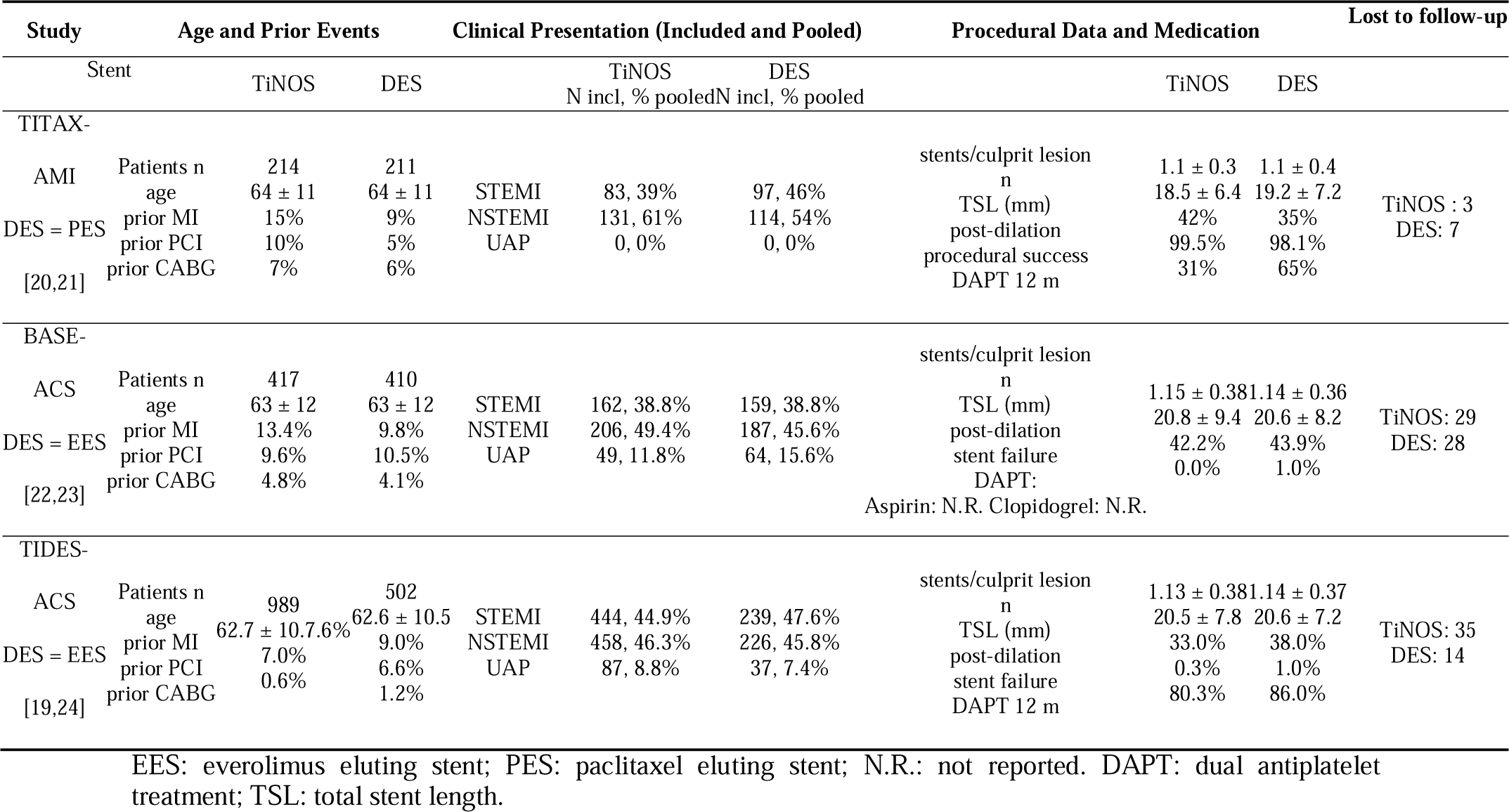
Eligible studies for 5-year follow-up. Baseline characteristics.

The previously reported number of patients by clinical presentation was inconsistent with the total sample size in the baseline characteristics of TIDES-ACS [19]. The authors provided the following *corrigendum* via P.K.: the need for: The number of patients with UAP is 87 in the TiNOS arm and 37 in the DES arm, not 126 and 61, respectively.

### 3.3. Individual Study Risk of Bias

The Cochrane instrument was used to rate individual study 5-year fol-low-up risk of bias. The only consistent risk of bias across the eligible RCTs was the operators’ knowledge of the stents they used (Figure 2).

**Figure 2.**
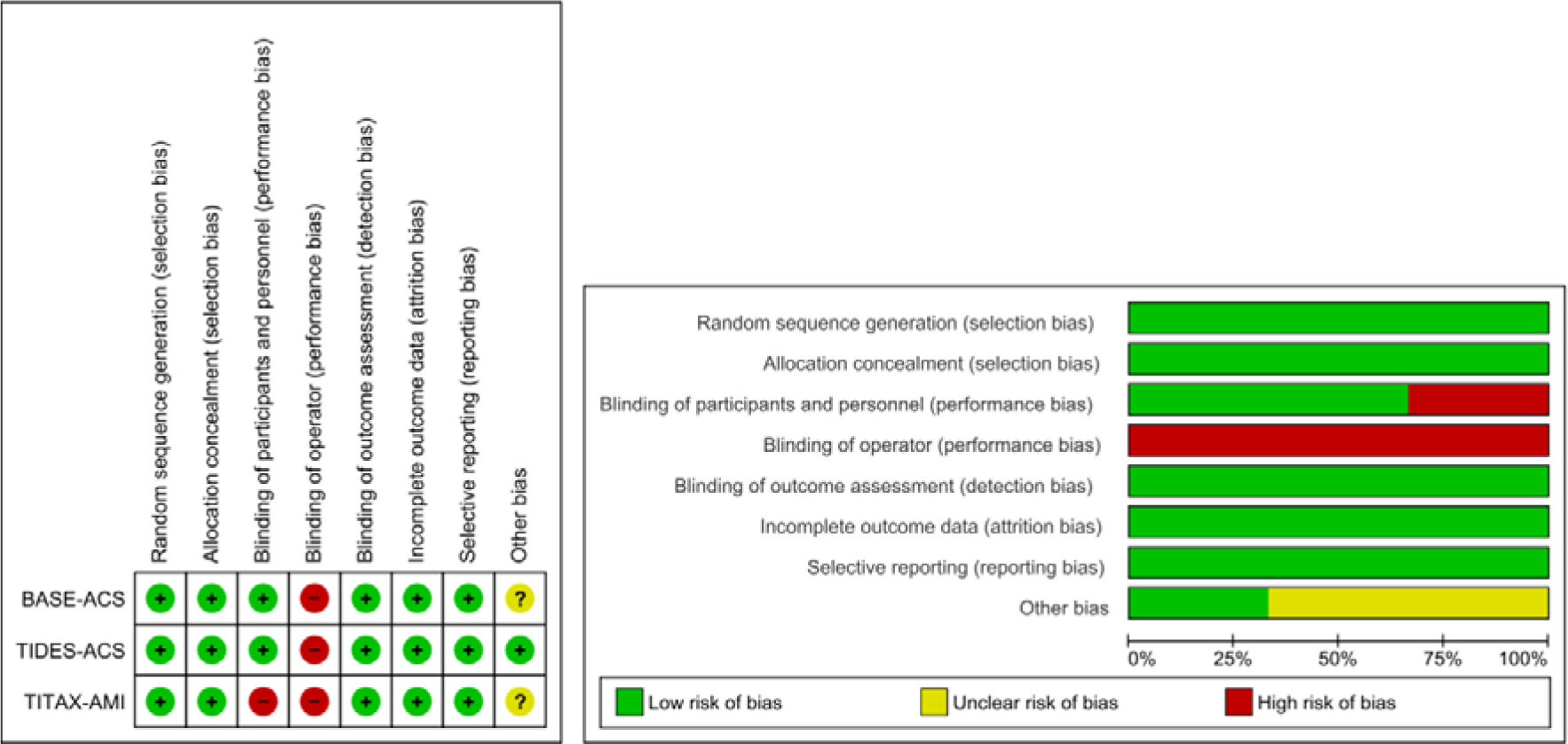
Individual Study Risk of Bias – 5-year follow-up.

### 3.4. Publication Bias Risk

The 5-year cumulative MACE RR funnel plot (Figure 3) detected no risk of publication bias, and the Harbord test (p = 0.08) detected no small-study effect.

**Figure 3.**
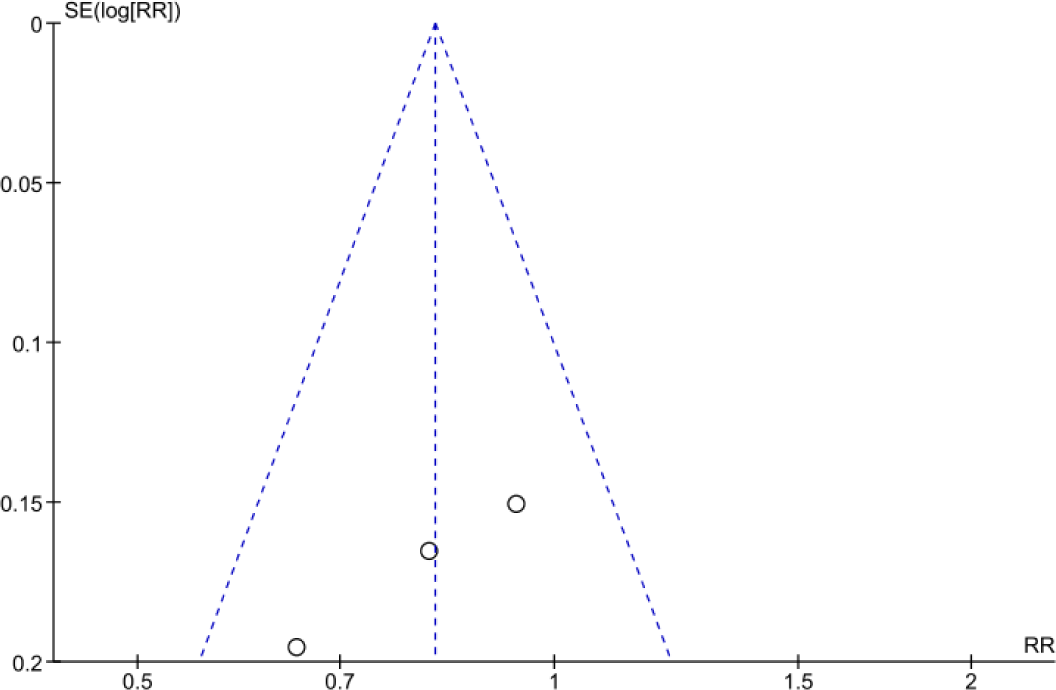
Funnel plot – Risk of Publication Bias – 5-year cumulative MACE RR.

### 3.5. Pooled Cumulative Outcomes

Figures 4 through 9 display the stratified pooled RRs of cumulative MACE, CD, MI, TLR, ST, and TD at 5-year follow-up. The polled RRs of MI, CD, and ST rates show a significantly lower event rate with TiNOS than with DES. The pooled RRs of TLR and MACE rates show TiNOS non-inferiority compared with DES, although the MACE rate with TiNOS is significantly lower than with DES.

**Figure 4.**
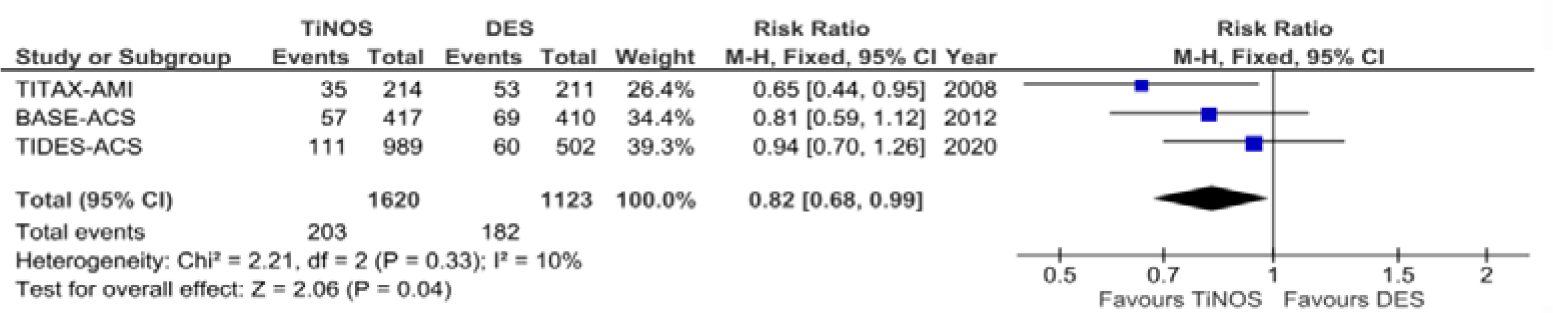
Forest plot – cumulative MACE – 5-year follow-up.

**Figure 5.**
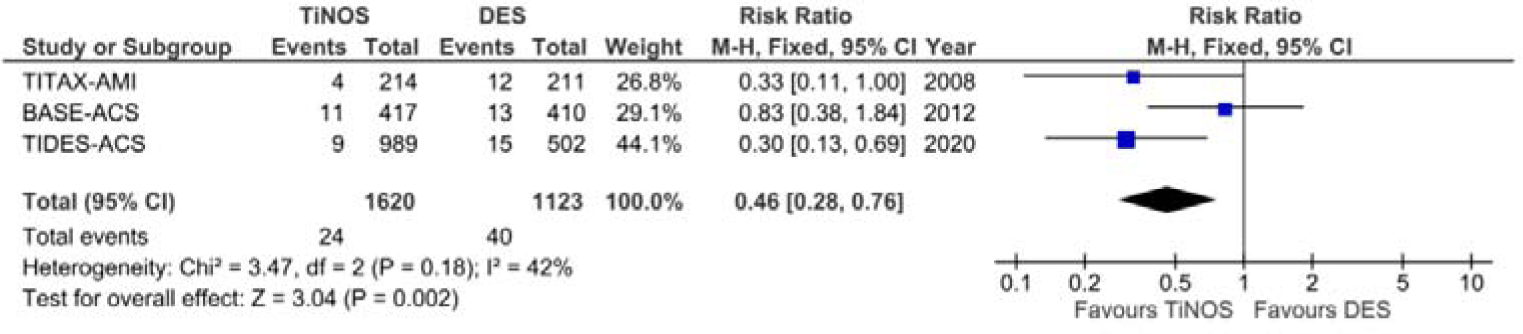
Forest plot – cumulative CD – 5-year follow-up.

**Figure 6.**
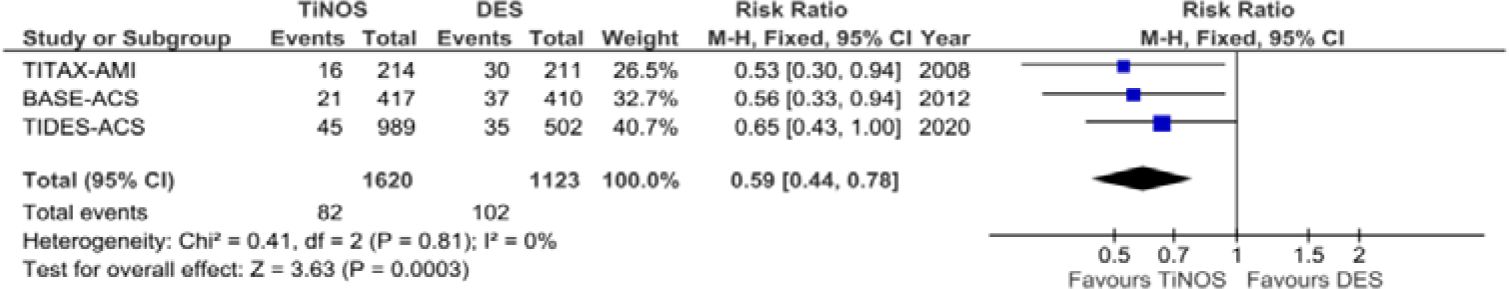
Forest plot – cumulative MI – 5-year follow-up.

**Figure 7.**
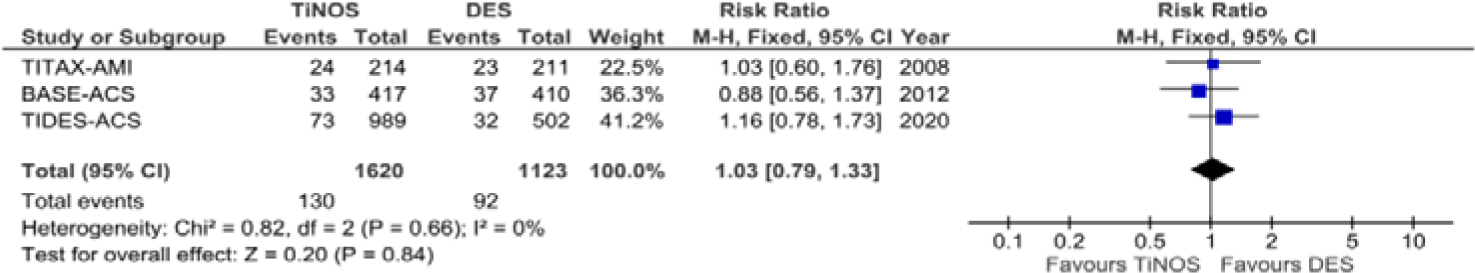
Forest plot – cumulative TLR – 5-year follow-up

**Figure 8.**
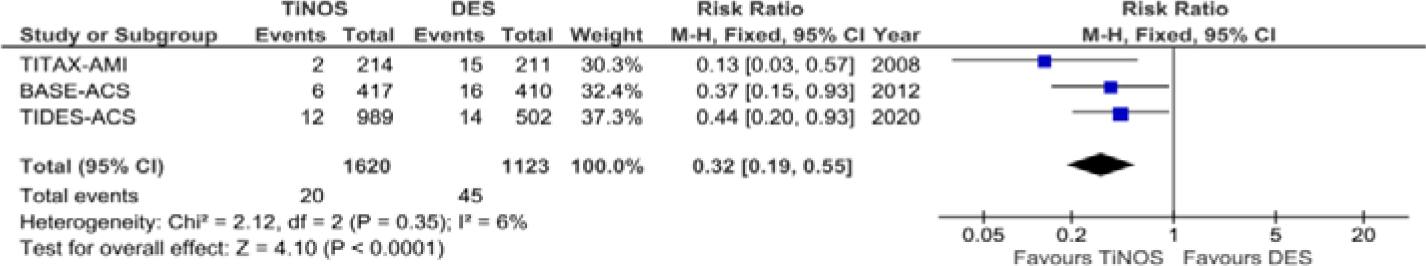
Forest plot – cumulative ST – 5-year follow-up

**Figure 9.**
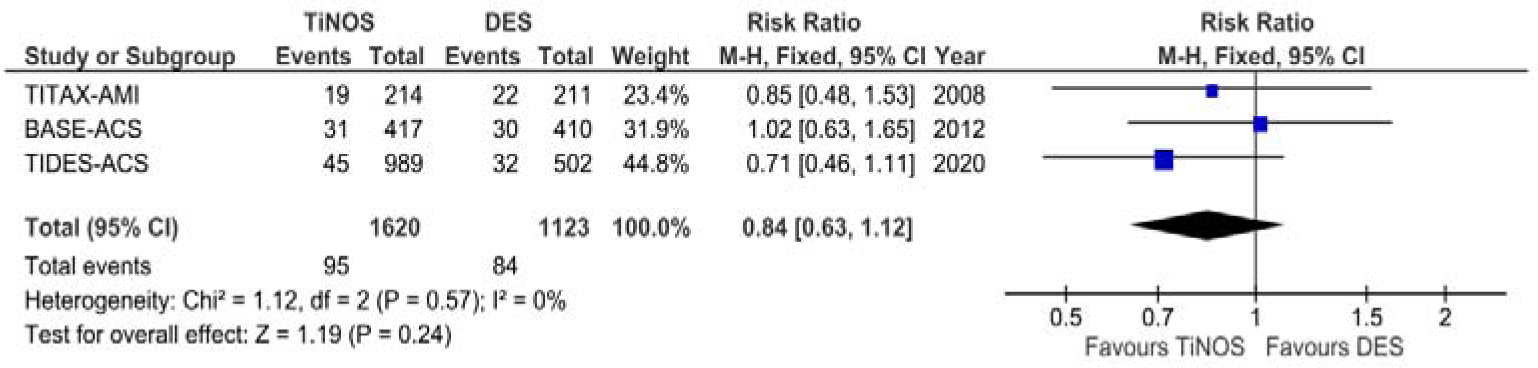
Forest plot – cumulative TD – 5-year follow-up

### 3.6. Sensitivity Analysis

The sensitivity analysis shows the impact on the pooled 5-year RRs of withdrawing one RCT at a time (Table 2). The pooled RR of all endpoints except MACE and CD are robust to sensitivity analysis. The GRADE rating shows high certainty levels with MACE, MI, and ST and moderate levels with TLR, CD, and TD.

**Table 2.** Sensitivity analysis of all endpoints TiNOS vs. DES at 5-year follow-up.

| M-H Fixed Effects RR and 95% CI after the Removal of: |  |  |  |  |  |  |
| --- | --- | --- | --- | --- | --- | --- |
| Endpoint | None | TITAX-AMI | TIDE | BASE-ACS | TIDES-ACS | Robustness |
| MACE | 0.82 [0.68, 0.99] | 0.88 [0.71, 1.09] | N.A. | 0.82 [0.65, 1.04] | 0.74 [0.58, 0.95] | No |
| CD | 0.46 [0.28, 0.76] | 0.51 [0.30, 0.89] | N.A. | 0.31 [0.16, 0.61] | 0.59 [0.31, 1.11] | No |
| MI | 0.59 [0.44, 0.78] | 0.61 [0.44, 0.85] | N.A. | 0.60 [0.43, 0.85] | 0.54 [0.37, 0.80] | Yes |
| TLR | 1.03 [0.79, 1.33] | 1.03 [0.76, 1.38] | N.A. | 1.11 [0.81, 1.54] | 0.94 [0.66, 1.32] | Yes |
| probable or definite ST | 0.32 [0.19, 0.55] | 0.40 [0.22, 0.73] | N.A. | 0.30 [0.15, 0.58] | 0.25 [0.12, 0.55] | Yes |
| TD | 0.84 [0.63, 1.12] | 0.84 [0.61, 1.16] | N.A. | 0.76 [0.54, 1.08]<sup>c</sup> | 1.03 [0.74, 1.45] | Yes |

N.A.: Not applicable

### 3.7. GRADE certainty of evidence

The GRADE rating (Table 3) shows high certainty levels with MACE, CD, MI, probable or definite ST, and moderate levels with TLR and TD.

**Table 3.**
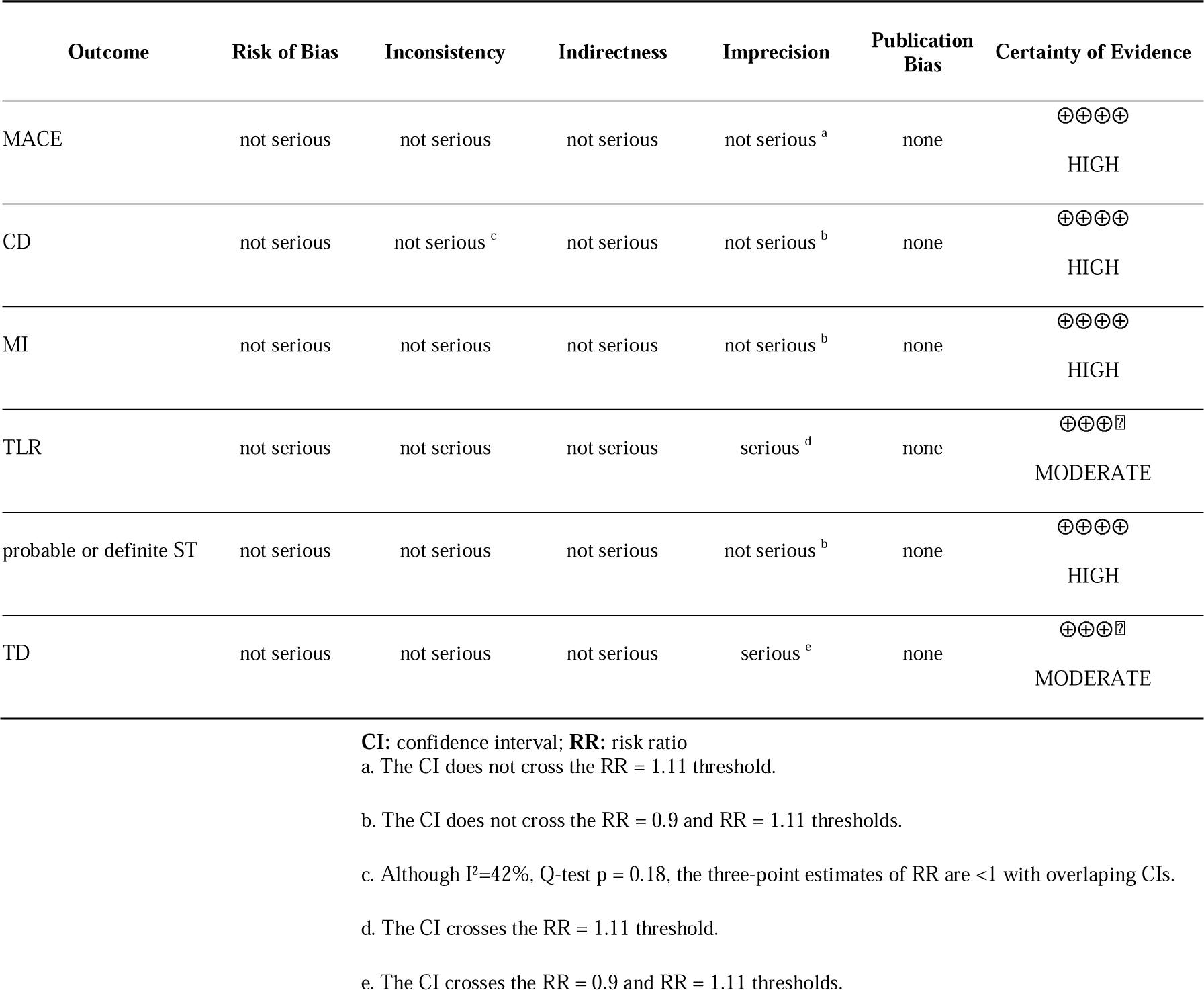
GRADE Summary of findings—TiNOS *vs*. DES in ACS at 5-year follow-up.

## 4. Discussion

### 4.1. Summary of the Results

The pooled analysis shows TiNOS’ non-inferiority at 5-year follow-up in MACE and superiority in MI, CD, and probable or definite ST, with a high level of certainty. It also shows the non-inferiority of TiNOS in TLR at a 5-year follow-up compared to a significantly higher rate of events than with DES at a 1-year follow-up, with a moderate certainty level due to the limited number of observations. TD’s RR is non-significant, but the certainty level is moderate for the same reasons as with TLR.

The increase in cumulative events in the two compared arms from one to five-year follow-ups and the updated precision grading method contribute to the increased certainty of outcomes, although RR estimates of some outcomes still have limited precision [12]. An extended follow-up of the patients beyond five years, if possible, would help identify further changes in all outcome variables, including TLR.

Overall, 5-year results show the long-term superior safety of TiNOS over DES in ACS with a high certainty level. The evidence also shows a progressive efficacy shift with the TLR rate that was significantly higher in TiNOS vs. DES at 1-year follow-up and reached non-inferiority at 5-year follow-up, although the number of observations remains insufficient for the level of evidence to become high. Extended follow-ups beyond five years should be monitored to check whether the shift continues over time.

The low rate of patients lost to follow-up in each RCT contributed to the continued low risk of bias at 5-year follow-up.

### 4.2. Clinical Validity

Assessing the clinical validity of the pooled RCTs requires checking that the 5-year MACE, CD, MI, TLR, probable or definite ST, and TD rates in ACS with TiNOS, EES, and PES are consistent with benchmark rates reported in larger samples of patients presenting with STEMI, NSTEMI, and UAP respectively. A PubMed and Cochrane database search for RCTs or me-ta-analyses of RCTs with those 5-year outcomes in ACS, STEMI, NSTEMI, or UAP returned no evidence with TiNOS but two meta-analyses comparing DES to BMS.

One individual patient data meta-analysis (IPDM) pooled data from 14 RCTs comparing DES vs. BMS in ACS [25]. Results were stratified outcome ratios up to 5-year follow-up in 34.5% of patients with CCS and 65.5% of patients with ACS. In the ACS subgroup, 7,739 patients were assigned to DES vs. 6,889 to BMS. The source publications of the included studies were reviewed, and ACS clinical presentations, DES types, outcomes definitions, sample sizes, and follow-up durations were checked [26–41]. None of the RCTs provide validation benchmarks for the DES arm.

One Cochrane review pooled 25 RCTs comparing the outcomes of DES vs. BMS in ACS [16]. The source publications were checked using the same methods as the IPDM [42–67]. Overall, 6,916 patients were treated with DES vs. 5,640 with BMS. A single RCT included patients with NSTEMI treated with EES, but the follow-up duration was only two years [57]. Another RCT included patients with STEMI treated with EES and reported outcomes at 5-year follow-up [42]. Another RCT included patients with STEMI treated with stainless-steel paclitaxel-eluting stents (PES) or BMS and reported outcomes at 5-year follow-up [63]. The latter two RCTs were the Cochrane review’s potential contribution to the validation benchmarks of the DES arm.

This meta-analysis comparing outcomes at five years with TiNOS vs. DES in ACS is the first of its kind. Therefore, clinical validation will require pooling additional future evidence to compare each stent and each clinical presentation with the corresponding benchmarks.

### 4.2. Generalization of the Results

Generalizing the results of this meta-analysis to a population is possible, either if the pooled study sample and the target population share comparable pre-PCI baseline characteristics or if the meta-analytic results can be standardized proportionately to the target population’s baseline characteristics. ACS covers the diversity of clinical presentations in emergency coronary care. Several epidemiological surveys have consistently shown that the incidence rate of STEMI decreases with age. In contrast, the incidence rate of NSTEMI increases with equal rates around 65 years of age, women present more frequently with NSTEMI or UAP than STEMI, and patients with STEMI are more often smokers than patients with NSTEMI [68–72]. Therefore, PCI outcomes at 5-year follow-up in any study should logically be associated with those base-line characteristics and other factors such as emergency care availability. A panel of epidemiological surveys on ACS hospital admissions and reperfusion rates shows significant differences in STEMI, NSTEMI, and UAP proportions across the sampled populations [68,69,72-76]. These differences show the difficulty of directly applying the outcomes of three pooled RCTs of this me-ta-analysis to populations with a different case mix. Reporting the pooled RRs stratified by clinical presentation can facilitate generalization to diverse populations through standardization.

## 5. Conclusions

The pooled outcomes at a five-year follow-up show that titani-um-nitride-oxide-coated stents are safer than drug-eluting stents in patients with acute coronary syndrome. The risk of cardiac death, myocardial infarction, and stent thrombosis is significantly lower with titanium-nitride-oxide-coated stents than with paclitaxel or everolimus-eluting metallic stents, with a high certainty level. The two stents display similar efficacy with non-significantly different target lesion revascularization rates with a moderate certainty level. All-cause mortality rates are not significantly different with an intermediate certainty level. Overall, the rates of device-oriented major adverse cardiac events are no significantly difference with a high certainty level. These pooled results are the first of their kind. Reporting relative risks stratified by clinical presentation would facilitate the clinical validation with external benchmarks and the generalization of the pooled results to patient populations with different proportions of acute coronary clinical presentations.

### Note

The ineligible records are references [2,16,66,77-122].

### Supplementary Materials

Not applicable.

### Author Contributions

F.D.: Designed the review protocol. F.D. and B.N.: Main reviewers and analysts. E.G.: Third reviewer and adjudicated disagreements. P.K.: Provided additional information about the methods of the individual trials and confirmed the number of patients lost to follow-up. All authors participated in finalizing the manuscript. All authors have read and agreed to the published version of the manuscript.

### Funding

This research received no external funding.

### Institutional Review Board Statement

Not applicable.

### Informed Consent Statement

Not applicable.

### Data Availability Statement

Not applicable. This work is a review of peer-reviewed articles available in Pubmed, Scopus, the Cochrane Library, and Web of Science.

## Appendix A. Detailed Search Strings in Each Database

### Pubmed search string

((bioactive OR (Titanium AND nitride AND oxide) OR TiNO OR TNO OR BAS) AND stent) AND (DES OR (drug AND eluting AND stent)) AND (RCT OR ((randomised OR randomised) AND controlled AND trial))

(“bioactivate”[All Fields] OR “bioactivated”[All Fields] OR “bioactivates”[All Fields] OR “bioactivating”[All Fields] OR “bioactivation”[All Fields] OR “bioacti-vations”[All Fields] OR “bioactive”[All Fields] OR “bioactives”[All Fields] OR “bioactivities”[All Fields] OR “bioactivity”[All Fields] OR ((“titanium”[MeSH Terms] OR “titanium”[All Fields] OR “titaniums”[All Fields]) AND (“nitridated”[All Fields] OR “nitridation”[All Fields] OR “nitride”[All Fields] OR “nitrided”[All Fields] OR “nitrides”[All Fields] OR “nitriding”[All Fields] OR “nitridized”[All Fields]) AND (“oxidability”[All Fields] OR “oxidable”[All Fields] OR “oxidant s”[All Fields] OR “oxidants”[Pharmacological Action] OR “oxidants”[MeSH Terms] OR “oxidants”[All Fields] OR “oxidant”[All Fields] OR “oxidate”[All Fields] OR “oxidated”[All Fields] OR “oxidates”[All Fields] OR “oxidating”[All Fields] OR “oxidation”[All Fields] OR “oxidations”[All Fields] OR “oxidative”[All Fields] OR “oxidatively”[All Fields] OR “oxidatives”[All Fields] OR “oxide s”[All Fields] OR “oxides”[MeSH Terms] OR “oxides”[All Fields] OR “oxide”[All Fields] OR “oxidic”[All Fields] OR “oxiding”[All Fields] OR “oxidisability”[All Fields] OR “oxidisable”[All Fields] OR “oxidisation”[All Fields] OR “oxidise”[All Fields] OR “oxidised”[All Fields] OR “oxidiser”[All Fields] OR “oxidisers”[All Fields] OR “oxidises”[All Fields] OR “oxidising”[All Fields] OR “oxidization”[All Fields] OR “oxidize”[All Fields] OR “oxidized”[All Fields] OR “oxidizer”[All Fields] OR “oxidizers”[All Fields] OR “oxidizes”[All Fields] OR “oxidizing”[All Fields])) OR “TiNO”[All Fields] OR “TNO”[All Fields] OR “BAS”[All Fields]) AND (“stent s”[All Fields] OR “stentings”[All Fields] OR “stents”[MeSH Terms] OR “stents”[All Fields] OR “stent”[All Fields] OR “stented”[All Fields] OR “stenting”[All Fields]) AND (“DES”[All Fields] OR (“drug”[All Fields] AND (“elutable”[All Fields] OR “elutant”[All Fields] OR “elute”[All Fields] OR “eluted”[All Fields] OR “elutent”[All Fields] OR “eluter”[All Fields] OR “eluters”[All Fields] OR “elutes”[All Fields] OR “eluting”[All Fields] OR “elution”[All Fields] OR “elutions”[All Fields]) AND (“stent s”[All Fields] OR “stentings”[All Fields] OR “stents”[MeSH Terms] OR “stents”[All Fields] OR “stent”[All Fields] OR “stented”[All Fields] OR “stenting”[All Fields]))) AND (“RCT”[All Fields] OR ((“random allocation”[MeSH Terms] OR (“random”[All Fields] AND “allocation”[All Fields]) OR “random allocation”[All Fields] OR “random”[All Fields] OR “randomization”[All Fields] OR “randomized”[All Fields] OR “randomisation”[All Fields] OR “randomisations”[All Fields] OR “randomise”[All Fields] OR “randomised”[All Fields] OR “randomising”[All Fields] OR “randomizations”[All Fields] OR “randomize”[All Fields] OR “randomizes”[All Fields] OR “randomizing”[All Fields] OR “randomness”[All Fields] OR “randoms”[All Fields] OR (“random allocation”[MeSH Terms] OR (“random”[All Fields] AND “allocation”[All Fields]) OR “random allocation”[All Fields] OR “random”[All Fields] OR “randomization”[All Fields] OR “randomized”[All Fields] OR “randomisation”[All Fields] OR “randomisations”[All Fields] OR “randomise”[All Fields] OR “randomised”[All Fields] OR “randomising”[All Fields] OR “randomizations”[All Fields] OR “randomize”[All Fields] OR “randomizes”[All Fields] OR “randomizing”[All Fields] OR “randomness”[All Fields] OR “randoms”[All Fields])) AND “controlled”[All Fields] AND (“clinical trials as topic”[MeSH Terms] OR (“clinical”[All Fields] AND “trials”[All Fields] AND “topic”[All Fields]) OR “clinical trials as topic”[All Fields] OR “trial”[All Fields] OR “trial s”[All Fields] OR “trialed”[All Fields] OR “trialing”[All Fields] OR “trials”[All Fields])))

Translations

bioactive: “bioactivate”[All Fields] OR “bioactivated”[All Fields] OR “bioacti-vates”[All Fields] OR “bioactivating”[All Fields] OR “bioactivation”[All Fields] OR “bioactivations”[All Fields] OR “bioactive”[All Fields] OR “bioactives”[All Fields] OR “bioactivities”[All Fields] OR “bioactivity”[All Fields]

Titanium: “titanium”[MeSH Terms] OR “titanium”[All Fields] OR “titani-um’s”[All Fields] OR “titaniums”[All Fields]

nitride: “nitridated”[All Fields] OR “nitridation”[All Fields] OR “nitride”[All Fields] OR “nitrided”[All Fields] OR “nitrides”[All Fields] OR “nitriding”[All Fields] OR “nitridized”[All Fields]

oxide: “oxidability”[All Fields] OR “oxidable”[All Fields] OR “oxidant’s”[All Fields] OR “oxidants”[Pharmacological Action] OR “oxidants”[MeSH Terms] OR “oxidants”[All Fields] OR “oxidant”[All Fields] OR “oxidate”[All Fields] OR “oxi-dated”[All Fields] OR “oxidates”[All Fields] OR “oxidating”[All Fields] OR “oxi-dation”[All Fields] OR “oxidations”[All Fields] OR “oxidative”[All Fields] OR “oxidatively”[All Fields] OR “oxidatives”[All Fields] OR “oxide’s”[All Fields] OR “oxides”[MeSH Terms] OR “oxides”[All Fields] OR “oxide”[All Fields] OR “oxi-dic”[All Fields] OR “oxiding”[All Fields] OR “oxidisability”[All Fields] OR “oxi-disable”[All Fields] OR “oxidisation”[All Fields] OR “oxidise”[All Fields] OR “oxi-dised”[All Fields] OR “oxidiser”[All Fields] OR “oxidisers”[All Fields] OR “oxidis-es”[All Fields] OR “oxidising”[All Fields] OR “oxidization”[All Fields] OR “oxi-dize”[All Fields] OR “oxidized”[All Fields] OR “oxidizer”[All Fields] OR “oxidiz-ers”[All Fields] OR “oxidizes”[All Fields] OR “oxidizing”[All Fields]

stent: “stent’s”[All Fields] OR “stentings”[All Fields] OR “stents”[MeSH Terms] OR “stents”[All Fields] OR “stent”[All Fields] OR “stented”[All Fields] OR “stenting”[All Fields]

eluting: “elutable”[All Fields] OR “elutant”[All Fields] OR “elute”[All Fields] OR “eluted”[All Fields] OR “elutent”[All Fields] OR “eluter”[All Fields] OR “eluters”[All Fields] OR “elutes”[All Fields] OR “eluting”[All Fields] OR “elu-tion”[All Fields] OR “elutions”[All Fields]

stent: “stent’s”[All Fields] OR “stentings”[All Fields] OR “stents”[MeSH Terms] OR “stents”[All Fields] OR “stent”[All Fields] OR “stented”[All Fields] OR “stenting”[All Fields]

randomized: “random allocation”[MeSH Terms] OR (“random”[All Fields] AND “allocation”[All Fields]) OR “random allocation”[All Fields] OR “random”[All Fields] OR “randomization”[All Fields] OR “randomized”[All Fields] OR “randomisation”[All Fields] OR “randomisations”[All Fields] OR “randomise”[All Fields] OR “randomised”[All Fields] OR “randomising”[All Fields] OR “randomi-zations”[All Fields] OR “randomize”[All Fields] OR “randomizes”[All Fields] OR “randomizing”[All Fields] OR “randomness”[All Fields] OR “randoms”[All Fields]

randomised: “random allocation”[MeSH Terms] OR (“random”[All Fields] AND “allocation”[All Fields]) OR “random allocation”[All Fields] OR “random”[All Fields] OR “randomization”[All Fields] OR “randomized”[All Fields] OR “randomisation”[All Fields] OR “randomisations”[All Fields] OR “randomise”[All Fields] OR “randomised”[All Fields] OR “randomising”[All Fields] OR “randomizations”[All Fields] OR “randomize”[All Fields] OR “randomizes”[All Fields] OR “randomizing”[All Fields] OR “randomness”[All Fields] OR “randoms”[All Fields]

trial: “clinical trials as topic”[MeSH Terms] OR (“clinical”[All Fields] AND “trials”[All Fields] AND “topic”[All Fields]) OR “clinical trials as topic”[All Fields] OR “trial”[All Fields] OR “trial’s”[All Fields] OR “trialed”[All Fields] OR “trial-ing”[All Fields] OR “trials”[All Fields]

### Cochrane database search string

((bioactive OR (Titanium AND nitride AND oxide) OR TiNO OR TNO OR BAS) AND stent) AND (DES OR (drug AND eluting AND stent)) AND (RCT OR ((randomised OR randomised) AND controlled AND trial)).

### Web of Science search string

(((bioactive OR (Titanium AND nitride AND oxide) OR TiNO OR TNO OR BAS) AND stent) AND (DES OR (drug AND eluting AND stent)) AND (RCT OR ((randomised OR randomised) AND controlled AND trial)))

### Embase search string

(((bioactive OR (Titanium AND nitride AND oxide) OR TiNO OR TNO OR BAS) AND stent) AND (DES OR (drug AND eluting AND stent)) AND (RCT OR ((randomised OR randomised) AND controlled AND trial)))/br.

## Data Availability

ll data produced are available online at https://pubmed.ncbi.nlm.nih.gov/

